# Cannabis use and risk of type 2 diabetes: a two-sample Mendelian randomization study

**DOI:** 10.1101/2020.09.21.20197343

**Authors:** Sebastian-Edgar Baumeister, Michael Nolde, Zoheir Alayash, Michael Leitzmann, Hansjörg Baurecht, Christa Meisinger

## Abstract

Cannabis has effects on the insulin/glucose metabolism. As the use of cannabis and the prevalence of type 2 diabetes increase worldwide, it is important to examine the effect of cannabis on the risk of diabetes. We conducted a Mendelian randomization study by using 19 single-nucleotide polymorphisms as instrumental variables for lifetime cannabis use and 14 SNPs to instrument cannabis use disorder, and linking these to type 2 diabetes risk using genome-wide association study data (lifetime cannabis use [N = 184,765]; cannabis use disorder [2,387 cases / 48,985 controls], type 2 diabetes [74,124 cases / 824 controls]). The MR analysis suggested no effect of lifetime cannabis use (inverse variance weighted odds ratio [95% confidence interval] = 1.00 [0.93-1.09], *P*-value = 0.935) and cannabis use disorder (OR = 1.03 [0.99-1.08]) on type 2 diabetes. Sensitivity analysis to assess potential pleiotropy led to no substantive change in the estimates. This study adds to the evidence base that cannabis use does not play a causal role in type 2 diabetes.

## Introduction

Cannabis is the most commonly used recreational drug globally (1). Diabetes has a worldwide prevalence of 8.8%, and by 2045 more than 600 million people are estimated to be living with diabetes (2). As more countries legalize the sale and consumption of cannabis, the number of users is continuing to rise (1). Given the increasing prevalence of both medicinal and recreational cannabis, it is increasingly important to understand its impact on public health. Although several observational studies have reported that cannabis use had favorable metabolic associations (3-5), including a lower prevalence of diabetes (3; 6), and lower glucose levels (7), evidence that cannabis is causally linked to the development of type 2 diabetes is insufficient. The current literature is limited by a preponderance of cross-sectional study designs (5). Available prospective observational studies (8) may be subject to social desirability and recall bias, and lack of valid cumulative exposure assessment (3; 9). Notably, cannabis users also engage in other behaviors that are associated with poor outcome. In particular, confounding bias (e.g., by tobacco smoking) may lead to spurious associations that preclude conclusions about causality. As cannabis smoking in combination with tobacco is the predominant method of use (10), due to positivity violations (11) (i.e., few cannabis users refrain from tobacco smoking), traditional confounding adjustment (e.g, regression, weighting, matching) is infeasible in observational studies. However, establishing causality is important, as this is essential for recommending public policies and clinical interventions. In this study, we use Mendelian randomization (MR) to examine whether cannabis use may lead to the development of type 2 diabetes. MR makes use of genetic instrumental variables to represent the exposure of interest and infers a relationship between exposure and outcome (12; 13). MR is not affected by reverse causation, as genetic variants are fixed at conception. MR is also less susceptible to environmental confounding compared with conventional observational studies because genetic instruments are assumed to affect the outcome only via the exposure and to be independent of confounders.

## Methods

### Data source

We performed two-sample, summary-based MR, in which the instrument-exposure and instrument-outcome associations were estimated in different samples. We retrieved associations of single nucleotide polymorphisms (SNPs) with lifetime cannabis use and cannabis use disorder from genome-wide association studies (GWAS) (14; 15). SNP-outcome associations were derived from two meta-analyses of genetic association studies of type 2 diabetes mellitus (16; 17).

### Selection of genetic instrumental variables for lifetime cannabis use and cannabis use disorder

GWAS summary statistics of 184,765 individuals of European descent for lifetime cannabis use, defined as any use during lifetime, were used (14). The data consisted of three sources and included the International Cannabis Consortium (ICC), 23andMe, and UK Biobank. Genotyping was performed on various genotyping platforms and standard quality control checks were performed before imputation. Genotype data were imputed using the 1000 Genomes phase 1 release reference set for ICC and 23andMe, and the Haplotype Reference Consortium reference set for the UK Biobank sample. Details regarding ethical approval and informed consent can be found in the original paper (14). A GWAS of diagnosed cannabis use disorder (ICD-10 F12.1– 12.9) (15) included 2,387 cases and 48,985 controls of the Lundbeck Foundation Initiative for Integrative Psychiatric Research (iPSYCH). Genotyping was performed using Illumina’s BeadArrays.

We selected independent 19 SNPs associated with lifetime cannabis use at *P* < 5 x 10^−7^ and independent 14 SNPs associated with cannabis use disorder at *P* < 5 x 10^−6^ using a PLINK clumping algorithm (r^2^ threshold = 0.01 and window size = 10mB) (Supplementary Table 1). We additionally adopted an approach (18; 19) to increase statistical power by lowering the *P*-values threshold (i.e., *P* < 5 x 10^−5^) for instrument selection and allowing for LD correlation (r^-2^ ≤ 0.3). This secondary approach provided 76 SNPs and 104 SNPs as instrumental variables for lifetime cannabis use and cannabis use disorder respectively.

**Table 1.** Mendelian randomization estimates for the relationship between cannabis use and type 2 diabetes mellitus

| Exposure | No. SNPs | Method | OR<sup>a</sup> | (95% CI) | P value |
| --- | --- | --- | --- | --- | --- |
| Lifetime cannabis use (Pasman et al. 2018) | 19 | Inverse variance weighted | 1.00 | (0.93;1.09) | 0.935 |
|  | 19 | Weighted median | 1.01 | (0.93;1.1) | 0.828 |
|  | 19 | Robust adjusted profile score | 1.01 | (0.93;1.08) | 0.888 |
|  | 19 | IVW radial | 1.00 | (0.93;1.09) | 0.935 |
|  | 19 | MR PRESSO | 1.00 | (0.93;1.09) | 0.936 |
|  | 76 | IVW (correlated variants) | 0.99 | (0.93;1.05) | 0.747 |
|  | 76 | Maximum likelihood (correlated variants) | 0.99 | (0.93;1.05) | 0.725 |
| Cannabis use disorder (Demontis et al. 2019) | 14 | Inverse variance weighted | 1.03 | (0.99;1.08) | 0.185 |
|  | 14 | Weighted median | 1.01 | (0.97;1.04) | 0.757 |
|  | 14 | Robust adjusted profile score | 1.02 | (0.98;1.06) | 0.317 |
|  | 14 | IVW radial | 1.03 | (0.99;1.08) | 0.185 |
|  | 12 | MR PRESSO | 1.01 | (0.99;1.03) | 0.422 |
|  | 104 | IVW (correlated variants) | 1.04 | (0.99;1.08) | 0.095 |
|  | 104 | Maximum likelihood (correlated variants) | 1.04 | (0.99;1.09) | 0.094 |

MR PRESSO, MR Pleiotropy RESidual Sum and Outlier. <sup>a</sup> OR (odds ratio) comparing cannabis use and non-use.

### GWAS summary statistics for diabetes

Genetic associations with type 2 diabetes were obtained from the GWAS summary statistics in the DIAGRAM consortium, which combined 32 GWAS results (Supplementary Table 2). The final GWAS was conducted in participants of European ancestry (cases□=□74,124, controls□=□824,006, 48.0% female) (16). We also used summary data from the second meta-analysis of type 2 diabetes GWAS (17) including 62,892 cases and 596,424 controls of European descent for a replication analysis.

### Patient consents, ethical approval and data availability

Written informed consent was obtained from each study participant. Study protocols for all cohorts were reviewed and approved by the appropriate institutional review boards. Detailed information on patient consent and ethics approval is available in the orginal GWAS (14-17). The summary statistics for the cannabis GWAS (14) are publicly available at https://www.ru.nl/bsi/research/group-pages/substance-use-addiction-food-saf/vm-saf/genetics/international-cannabis-consortium-icc/ (access date: 2020/07/20). The cannabis use disorder data are available at https://ipsych.dk/en/research/downloads/data-download-agreement-ipsych-secondary-phenotypes-cannabis-2019/ (access date: 2020/07/20). The primary type 2 diabetes GWAS (16) summary data are available at http://diagram-consortium.org/downloads.html (access date: 2020/07/20) and the summary data from the second type 2 diabetes GWAS (17) is available at https://cnsgenomics.com/content/data (access date: 2020/07/20). The study adhered to the MR-Strobe statement.

### Statistical power

The a priori statistical power was calculated according to Brion et al. (20). The 19 SNPs for lifetime cannabis use explained 0.06% of the phenotypic variance. Given a type 1 error of 5%, we had sufficient statistical power (≥ 80%) when the expected odds ratio (OR) for type 2 diabetes was ≤ 0.65 in genetically instrumented cannabis use. In the secondary approach (18) using 104 weak, correlated variants for lifetime cannabis use, we achieved a power of 81% to detect an OR of 0.77.

### Statistical analyses

Harmonization was performed to rule out strand mismatches and to ensure alignment of effect sizes. Wald ratios were calculated by dividing the per-allele log OR for each SNP from the diabetes GWAS by the corresponding log OR of the same SNP in the cannabis GWAS. We estimated the effect of cannabis use on diabetes risk by performing a multiplicative random effects inverse-variance weighted (IVW) meta-analysis of Wald ratios(19). IVW-results are presented as mean OR for type 2 diabetes comparing cannabis use and non-use. MR is based on the assumption that SNP-outcome effects are mediated solely through the exposure (12; 21). Violations of this assumption through horizontal pleiotropy, whereby the instruments exert an effect on the outcome independent of the exposure can introduce bias. To examine possible violations of this assumption, we checked each candidate SNP and its proxies (r^2^>0.8) in PhenoScanner (22) for previously reported associations (P<5×10^−8^) with confounders, and performed additional analyses excluding potentially pleiotropic SNPs. Of the risk factors judged to have sufficient evidence of relationship with type 2 diabetes (23), we considered alcohol drinking and tobacco smoking to be the only common causes of cannabis use and type 2 diabetes (24). Furthermore, statistical sensitivity analyses more robust to the inclusion of potentially pleiotropic variants can be used to help establish the validity of causal inference from MR analysis. Valid genetic instruments should furnish similar estimates of effect (21). This can be assessed using a modified Q and the I^2^ statistic (21). If Q and I^2^ detect heterogeneity among ratio estimates, this points to pleiotropy. If the pleiotropy is ‘balanced’ (i.e., pleiotropic effects are independent in the magnitude of the SNP-exposure associations; and if the mean pleiotropic effect is zero), the effect can be reliably estimated by the multiplicative random effects IVW method (19; 21). However, if the mean pleiotropic effect is nonzero, as shown by the presence of a deviation from a zero intercept of an MR Egger regression (12), robust meta-analytic methods are indicated (21; 25). Classes of robust methods each relying on different sets of assumptions can assist in protecting against pleiotropy. We followed Slob and Burgess (25) to report estimates from at least one method of three classes of robust methods: (1) consensus class (weighted median (12)), (2) outlier robust class (MR-Pleiotropy Residual Sum and Outlier (MR-PRESSO) (25), Radial regression (26)), (3) modeling methods class (Robust Adjusted Profile Score (25)). We also performed a leave-one-out analysis to assess whether the IVW estimate was driven or biased by a single SNP and replicated the analysis using summary data from the second diabetes GWAS (17). Furthermore, we used the multiplicative random effects IVW and maximum likelihood methods for correlated instrumental variables (18). Analyses were performed using the meta (4.11.0), TwoSampleMR (0.5.5) (27), MendelianRandomization (0.4.3) and MRPRESSO (1.0) packages in R (version 4.0.2).

## Results

The ORs from the IVW models for the effect of cannabis use and cannabis use disorder on type 2 diabetes were 1.00 (95% CI: 0.93-1.09, *P*-Value = 0.935) and 1.03 (95% CI: 0.99-1.08, *P*-Value = 0.935), respectively (Table 1). None of the instrumental variables was associated with tobacco smoking or alcohol consumption in published GWAS in PhenoScanner (Supplementary Table 3). There was moderate heterogeneity between Wald ratios in the IVW estimates of lifetime cannabis use and cannabis use disorder (Supplementary Table 4). The intercept from the MR Egger regression was not different from zero (Supplementary Table 5). Estimates were similar when using models more robust to directional pleiotropy (Tables 1). Leave-one-out analyses revealed that no single SNP drove the results (Supplementary Table 5). When summary data from the second diabetes GWAS meta-analysis was used, the IVW ORs were 0.94 (95% CI: 0.86-1.04, *P*-Value = 0.217) and 1.04 (95% CI: 0.86-1.04, *P*-Value= 0.217) for lifetime cannabis use and cannabis use disorder, respectively (Supplementary Table 6). Estimates were similar when correlated, weak instrumental variables were used (Table 1).

## Discussion

Using genetic instrumental variables for lifetime cannabis use from GWAS of more than 180,000 individuals and 74,000 cases of diabetes, we examined the relationship between lifetime cannabis use and type 2 diabetes. We additionally used cannabis use disorder as an exposure variable that reflects heavy cannabis use. This study provides no evidence for a role of cannabis use in the development of type 2 diabetes. Several cross-sectional studies have suggested that cannabis has beneficial metabolic effects (3; 5; 7; 28-30) but prospective observational studies have not supported inverse associations with type 2 diabetes (8). Cannabis intake stimulates appetite and increases the use of low nutritional value carbohydrates and it promotes adipogenesis, which is expected to increase insulin resistance (4; 31). Consequently, the Coronary Artery Risk Development in Young Adults (CARDIA) (8) cohort found an increased risk of pre-diabetes among current (OR: 1.65, 95% CI: 1.15-2.38) and lifetime cannabis users (OR: 1.49, 95% CI: 1.06-2.11), but no increased risk for manifest diabetes. Another prospective study of more than 17,000 Swedish men and women found no association between lifetime cannabis use and diabetes risk (32). A small randomized double-blind trial (33) showed that in patients with type 2 diabetes, tetrahydrocannabivarin (a CB_1_ receptor antagonist at low dose) decreased fasting plasma glucose levels and improved pancreatic β-cell function. Likewise, in another small randomized trial among healthy cannabis users (34), cannabis use lowered blood insulin, glucagon-like peptide 1, and ghrelin levels.

However, despite speculation about the potential metabolic effect of cannabinoids (31; 35), previous observational research has mostly applied cross-sectional designs and did not establish dose-response associations. Furthermore, existing observational research may have been subject to unaccounted confounding (cigarette smoking), reverse causation (people who feel unwell and are at risk quit or cannot tolerate cannabis (4)), and measurement error (erroneous recall or social desirability).

Notable strengths of the present study are the large sample size of the outcome GWAS meta-analyses that enabled considerable statistical precision. Sensitivity analyses identified that the estimates from MR were robust to various approaches that tested for stability and model violations. A limiting factor is that the instrumenting SNPs explained little phenotypic variance. However, our study had sufficient statistical power to detect the previous observationally reported effect sizes for cannabis use and diabetes (5; 31), and all F statistics (where F<10 indicates weak instrument bias) suggested valid instruments. When larger cannabis GWAS will become available, this will help to identify additional SNPs that could serve as instruments and improve the precision of our MR estimates. The present study did not allow us to investigate the route of administration, the composition of plant components, or the age at exposure to cannabis. The MR models employed assumed no interaction (e.g., gene-environment), and a linear relationship between cannabis use and type 2 diabetes. In summary, our findings provide support for a lack of a causal association between cannabis use and the risk of type 2 diabetes.

## Data Availability

The summary statistics for the cannabis GWAS (14) are available at https://www.ru.nl/bsi/research/group-pages/substance-use-addiction-food-saf/vm-saf/genetics/international-cannabis-consortium-icc/ (access date: 2020/07/20). Thr cannabis use disorder data are available at https://ipsych.dk/en/research/downloads/data-download-agreement-ipsych-secondary-phenotypes-cannabis-2019/ (access date: 2020/07/20). The primary type 2 diabetes GWAS (16) summary data are available at http://diagram-consortium.org/downloads.html (access date: 2020/07/20) and the summary data from the second type 2 diabetes GWAS (17) is available from https://cnsgenomics.com/content/data (access date: 2020/07/20).

## Acknowledgements

The authors acknowledge and thank the investigators of the original studies (14-17) for sharing the cannabis and type 2 diabetes GWAS data used in this study.

## Author Contributions

Conception and design: Sebastian E Baumeister, Zoheir Alayash, Michael Nolde, Christa Meisinger. Development of methodology: Sebastian E Baumeister, Zoheir Alayash, Michael Nolde, Hansjörg Baurecht Acquisition of data (provided animals, acquired and managed patients,provided facilities, etc.): Sebastian E Baumeister, Zoheir Alayash. Analysis and interpretation of data (e.g., statistical analysis, biostatistics,computational analysis): Sebastian E Baumeister, Zoheir Alayash, Michael Nolde, Hansjörg Baurecht. Writing, review, and/or revision of the manuscript: Sebastian E Baumeister, Zoheir Alayash, Michael Leitzmann, Hansjörg Baurecht, Christa Meisinger. Administrative, technical, or material support (i.e., reporting or organizing data, constructing databases): Zoheir Alayash, Michael Nolde.

